# The structural, functional, and neurophysiological connectome of mild traumatic brain injury: a DTI, fMRI and MEG multimodal clustering and data fusion study

**DOI:** 10.1101/2024.06.24.24309379

**Authors:** Jing Zhang, Kevin Solar, Kristina Safar, Rouzbeh Zamyadi, Marlee M. Vandewouw, Leodante Da Costa, Shawn G. Rhind, Rakesh Jetly, Benjamin T. Dunkley

**Affiliations:** Defence Research and Development Canada, Toronto, Canada; Neurosciences & Mental Health, Hospital for Sick Children Research Institute, Toronto ON, Canada; Institute of Biomedical Engineering, University of Toronto, Toronto, Canada; Autism Research Centre, Holland Bloorview Kids Rehabilitation Hospital, Toronto, Canada; Sunnybrook Health Sciences Centre, Toronto, ON, Canada; Department of Surgery, University of Toronto, Toronto ON, Canada; Faculty of Kinesiology and Physical Education, University of Toronto, Toronto ON, Canada; Institute of Mental Health Research (IMHR), University of Ottawa, Ottawa ON, Canada; Diagnostic & Interventional Radiology, Hospital for Sick Children, Toronto ON, Canada; Department of Medical Imaging, University of Toronto, Toronto ON, Canada; Department of Psychology, University of Nottingham, Nottingham, UK

**Keywords:** Mild traumatic brain injury, concussion, magnetoencephalography, functional magnetic resonance imaging (fMRI), diffusion tensor imaging, machine learning, data fusion

## Abstract

The clinical presentation and neurobiology of mild traumatic brain injury (mTBI) - also referred to as concussion - are complex and multifaceted, and interrelationships between neurobiological measures derived from neuroimaging are poorly understood. This study applied machine learning (ML) to multimodal whole-brain functional connectomes from magnetoencephalography (MEG) and functional magnetic resonance imaging (fMRI), and structural connectomes from diffusion tensor imaging (DTI) in a test of discriminative accuracy in cases of mTBI. Resting state MEG (amplitude envelope correlations), fMRI (BOLD correlations), and DTI (fractional anisotropy, FA; streamline count, SC) connectome data was acquired in 26 controls without mTBI (all male; 27.6 ± 4.7 years) and 24 participants with mTBI (all male; 29.7 ± 6.7 years) in the acute-subacute phase of injury. ML with data fusion was used to optimally identify modalities and brain features for discriminating individuals with mTBI from those without. Univariate group differences were only found for MEG functional connectivity, while no differences were found for fMRI or DTI. Functional connectivity (fMRI and MEG) showed robust unimodal classification accuracy for mTBI, followed by structural connectivity (DTI), where FA showed marginally better classification performance than SC, but SC outperformed FA in data interpretation and fusion. Perfect, unsupervised separation of participants with and without mTBI was achieved through participant fusion maps featuring all three data modalities. Finally, the MEG-only full feature fusion map showed group differences, and this effect was eliminated upon integrating DTI and fMRI datasets. The markers identified here align well with prior multimodal findings in concussion and highlight modality-specific considerations for their use in understanding network abnormalities of mTBI.

## Introduction

Mild traumatic brain injury (mTBI), also known as concussion, is a neurological injury that causes a range of metabolic, microstructural, and behavioral effects, which often lead to a reduced quality of life [1], [2]. mTBI is common – the annual incidence rates in Ontario, Canada alone for diagnosed concussion is 1.2% of the population [3] - yet our understanding of the neurobiological mechanisms underlying the symptoms and protracted recovery is limited. The clinical presentation of mTBI varies greatly between individuals; consequently, there is no single pathognomonic test to diagnose mTBI [4], [5], [6]. Given the high prevalence of mTBI and the associated burden it places on the healthcare system and society, it is important to identify neuropathological characteristics and develop objective indices for accurate diagnosis and prognosis.

Neuroimaging has revealed indicators of mTBI pathophysiology, including altered neuronal activity and microstructure, but results are inconsistent [7], [8]. Multimodal neuroimaging studies are useful because they provide complementary information about structural and functional brain circuits [9], [10], [11]. However, interpretation and integration of multimodal neuroimaging data remains challenging due to the heterogeneity of the neurobiological signals measured across imaging modalities [12] and the complexity of the data [13].

One approach that has attempted to fuse imaging data is through *connectomics*. Connectomics is the study of structural and functional brain networks, including the segregation and integration of brain regions that allow for daily functioning [14], [15]. In brain networks, nodes represent brain regions and edges signify the connections between regions [16], [17]. Structural connectivity is defined by the physical connections between given brain regions, and can be quantified by various metrics, including those derived from diffusion weighted imaging (DWI), which produces parameters that characterize tissue microstructure (e.g., axonal packing, myelination), including streamline count (SC) and fractional anisotropy (FA), while functional connectivity refers to the statistical correlation of brain activity between regions, such as that derived from functional MRI (fMRI) time series data. Importantly, connectomics allows for multiple imaging modalities to be expressed similarly (i.e., whole-brain structural and functional connectivity can be calculated between the same anatomical regions across multiple types of neuroimaging data), allowing comparison of results within and across modalities.

Connectomics have identified extensive structural [18], [19], [20] and functional network dysfunction in mTBI [19], [21]. DWI is one way to commonly used to quantify structural connectivity. Functional connectivity is generally measured by fMRI, and less frequently by electrophysiological tools such as magnetoencephalography (MEG). The basis of fMRI connectivity is the blood oxygen level dependent (BOLD) response, the covariation of which between brain regions defines functional connectivity [22]. MEG measures the magnetic fields produced by neuronal activity [23] and can be used to quantify functional connectivity in multiple ways, and the most common approach is amplitude envelope correlations (AEC), a statistical measure of association between fluctuating oscillatory activity in each brain region. Notably, MEG AEC correlates highly with BOLD resting state fMRI [24].

Each of these modalities individually and in various combinations have revealed altered brain connectivity profiles in mTBI [25], [26], [27], [28], [29], [30]. The rich variety of information available in multimodal connectomics presents a way to understand multifaceted dysregulation to brain connectivity that occurs after injury. Assessing which neuroimaging modality or combination of modalities - DTI, fMRI, and/or MEG - is optimally sensitive to and best captures network dysfunction in mTBI is valuable, as well as exploring more advanced computational approaches to integrate complex neuroimaging datasets.

Recent advances in artificial intelligence (AI)-driven high-throughput informatics, as well as high-performance computing, it is now possible to apply machine learning (ML) methodology to multimodal neuroimaging [31]. ML has been extensively used for feature selection and classification modelling in neuropathology, with a dynamic combination of the ML-driven feature selection and classification modelling driving both basic science and clinical research advances [32]. For example, a DTI study demonstrated promising classification performance for mTBI by utilizing multiple ML methods, including support vector machine (SVM) and random forest (RF) [33] with similar methods successfully applied to fMRI data [34]. Using an in-house developed ML feature selection and classification pipeline [35], several studies from our group have demonstrated optimal performance for mTBI classification using MEG, which identified key neural signatures underlying mTBI-linked neuropathophysiology [36], [37], [38] and allowed for differential classification of post-traumatic stress disorder (PTSD) [37].

Multimodal neuroimaging fusion analysis [39] could provide a step-change in understanding connectome level changes in mTBI [10], having already been applied in various neuropsychiatric disorders [40]. Generally, multimodal integration includes *fusion* and *correlation* aspects. Multimodal *fusion* approaches could afford insights into neuropathology beyond unimodal investigations and improve sensitivity and specificity in disease [41]. For example, network similarity-based data fusion using similarity network fusion (SNF) has enabled sub-group identification, a crucial step towards personalized precision medicine [42]. To date, multimodal neuroimaging *fusion* has not been applied in mTBI research.

Given the breadth of information available across different neuroimaging techniques, the potential in ML-powered data mining, and the lack of an objective diagnostic/prognostic tools for mTBI, ML-assisted multimodal data integration could significantly improve our understanding of a heterogenous condition. The aim of this study was to examine the capacity for ML-assisted multimodal neuroimaging integration to discriminate mTBI status. First, within-modality connectomics with ML was applied to structural (DTI) and functional connectivity (fMRI, MEG). Second, a multimodal integration analysis of DTI, fMRI, and MEG datasets was conducted using the SNF fusion procedure [43]. The goals of the integration analysis were to define modality combination and feature identification for optimal participant group separation.

## Methods and Materials

### Participants

Fifty participants were recruited, stratified into two groups: controls (all male, n = 26; mean age = 27.6 ± 4.7 years, range = 20-39 years) and mTBI (all male, n = 24; mean age = 29.7 ± 6.7 years, range = 20-44 years). Participants in the mTBI group were recruited through the Sunnybrook Health Science Center in Toronto, Canada, after admittance to the Emergency Department for their first mild traumatic brain injury, with a diagnosis determined by a physician specializing in head trauma. All participants were scanned within 3 months (<90 days) of injury, with an average of ∼30 days since injury.

For both groups, inclusion criteria included: English-speaking, able to understand instructions, and able to give informed consent. Exclusion criteria included any MRI or MEG contraindications, implanted medical devices, a diagnosed history of pre-trauma seizures or other neurological or psychiatric disorders, active substance abuse, and certain ongoing medications (i.e., anticonvulsants, benzodiazepines, and/or GABA antagonists) known to directly or significantly influence neural oscillations. All subjects had an MRI scan reviewed by a neuroradiologist and no positive clinical indications were found. The study was approved by Sunnybrook and the Hospital for Sick Children Research Ethics Boards.

### Image Processing Magnetoencephalography (MEG)

Resting state MEG data were acquired in a supine position using a CTF 151 channel system at the Hospital for Sick Children. This data has previously been published [36] – the DTI and fMRI data have not been published previously. Briefly, fiducial coils were attached to the nasion and left and right preauricular points to allow for continuous monitoring of head motion. Data were collected at a sampling rate of 600 Hz, bandpass filtered offline using a high-pass filter of 1 Hz and a low-pass of 150 Hz, with a 60 Hz notch filter and subsequently analyzed in Fieldtrip Toolbox [44], with virtual electrodes derived using a beamformer approach from 90 Automated Anatomical Labelling (AAL) Atlas areas – the same atlas was used for defining parcellations in DTI and fMRI analyses. Further details are described in the **Supplementary Methods.**

### Magnetic Resonance Imaging

T1-weighted images, DTI, and resting state fMRI data were acquired using a Siemens Trio 3T scanner with a 12-channel head coil at the Hospital for Sick Children. Anatomical T1-weighted images were acquired using a SAG-MPRAGE sequence (TR = 2.3 s, TE = 2.96 ms, flip angle = 9°, FOV = 240 x 256 mm^2^), followed by DTI using a spin echo EPI acquisition sequence (60 directions at b = 1000 s/mm^2^, TR = 8.8 s, TE = 87 ms, FOV = 244 × 244 mm^2^, 2 x 2 x 2 mm^3^), and finally fMRI (TR = 2.3 s, TE = 30 ms, flip angle = 70). Details on processing can be found in **Supplementary Methods.**

### Statistical Analysis

#### False Discovery Rate (FDR) analysis

Independent t-tests determined between-group edgewise differences in structural (DTI) and functional connectome (fMRI, MEG) data, while controlling for age. The p-value was estimated using non-parametric permutation testing (100,000 permutations) with false discovery rate (FDR) correction for multiple comparisons, with significance held at *p* < 0.025 [45], [46]. The Network Based Statistics (NBS) toolbox was used to compute the analyses using the FDR option (https://www.nitrc.org/projects/nbs).

### Machine learning and multimodal data integration analysis

**Figure 1.**
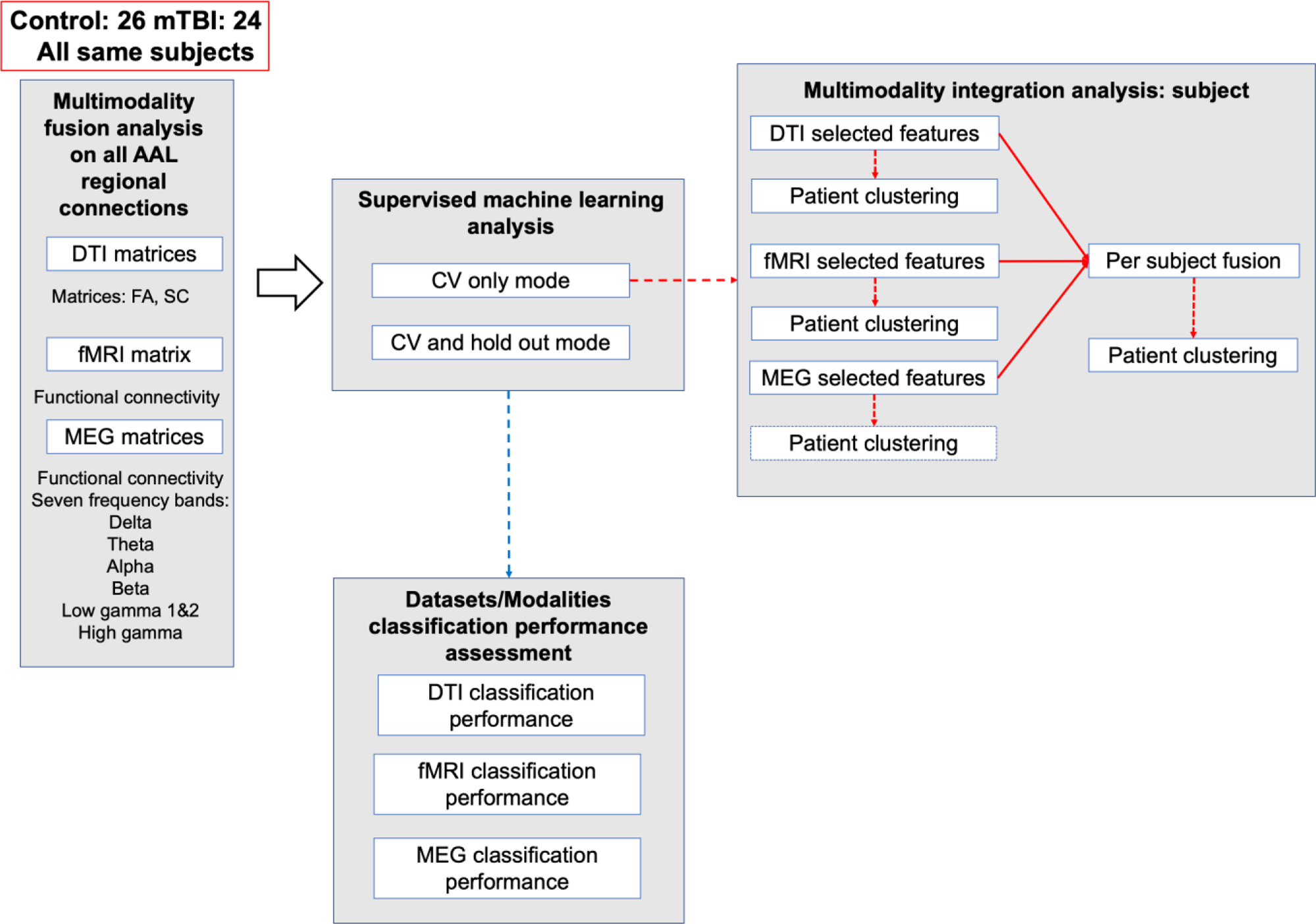
The complete workflow for multimodal data integration analysis. The focus here is to dynamically combine ML feature selection and modelling with the KNN (k-nearest neighbour) network-based SNF analysis [42]. Our multimodal integration workflow was a dynamic combination of supervised and unsupervised ML processes.

### Overall workflow

Input data consisted of connectivity matrices from the three modalities (i.e., DTI, fMRI, and MEG). DTI and MEG generated multiple data types - DTI included FA (fractional anisotropy) and SC (streamline count) weighted structural connectomes, and MEG featured seven canonical frequency bands (**Figure 1**). For MEG, we focused on the frequencies exhibiting the best ML classification performances, identifying frequencies with the optimal receiver operator characteristic-area under curve (ROC-AUC) values (>0.8) from both modelling modes and with the least or no overfitting, by comparing training and holdout test data ROC-AUC values.

The multimodal fusion analysis was conducted for participants with the objective to assess group separation and participant clustering. Data modality combinations for fusion were determined based on the group separation performance (according to unsupervised hierarchical clustering) with single modal datasets. DTI networks were quantified either by FA or SC datasets, and all the fusion networks contained either none, or one of the DTI (FA or SC) data sets, not both.

### Multimodal analysis with subject similarity fusion

For the subject similarity fusion, the fusion networks were ultimately based on the ML selected features for each data modality. The subject similarity fusion was examined if the subjects would be separated into the subject groups in an unsupervised manner (hierarchical clustering, “Ward.D2” method) [47], using the information from the three imaging techniques. Moreover, we investigated which data modality, or combination of modalities, led to the best subject group clustering. Additionally, the subject similarity fusion also revealed potential similar subject sub-groups within their respective subject groups. To identify the best fusion network(s) for subject group clustering, we started with the single modality KNN networks. Based on the subject clustering performance, the best single modal dataset was used as the “seed” dataset, to which other datasets would be added to form fusion networks. Detailed fusion analysis methods can be viewed in **Supplementary Methods**.

### Supervised machine learning workflow

The core supervised ML workflow followed our previous reports [35], [37], [38], with modifications. Briefly, a 10-fold cross validation (CV) process was used for both feature selection and classification modelling. A detailed description of the ML workflow can be viewed in **Supplementary Methods.**

BrainNet Viewer [48] was used to visualize the features selection (FS) results for the datasets used for the follow-up multimodal analysis. Principal component analysis (PCA) was used as an unsupervised clustering visualization method to communicate the capability of selected features in separating the two subject groups, for the single modal datasets. For PCA plots, the first three PCs (principal components) were visualized.

## Results

### MEG reveals between groups differences in connectivity not evident in DTI or fMRI

The mTBI group exhibited significantly lower MEG functional connectivity in the alpha, beta, and low gamma bands. In the alpha and beta band, AEC was significantly decreased in mTBI compared to controls (*p_corr_* < 0.025; **Figure 2**) among distributed brain areas, including cortical and subcortical regions, including the bilateral thalamus, pallidum, and caudate nucleus, with connections to other subcortical, limbic, as well as parietal, occipital and frontal areas. Decreased AEC in the low gamma band in mTBI was observed in the in the right hippocampus and right lingual gyrus, and the left superior frontal gyrus, medial and the right angular gyrus (*p_corr_* < 0.025; **Figure 2**). No group differences were found in the delta, theta, low gamma 2 or high gamma frequency bands, or for SC, FA or fMRI connectivity after correction for multiple comparisons (*p_corr_* < 0.025; nor at a less stringent significance threshold, *p_corr_* < 0.05).

**Figure 2.**
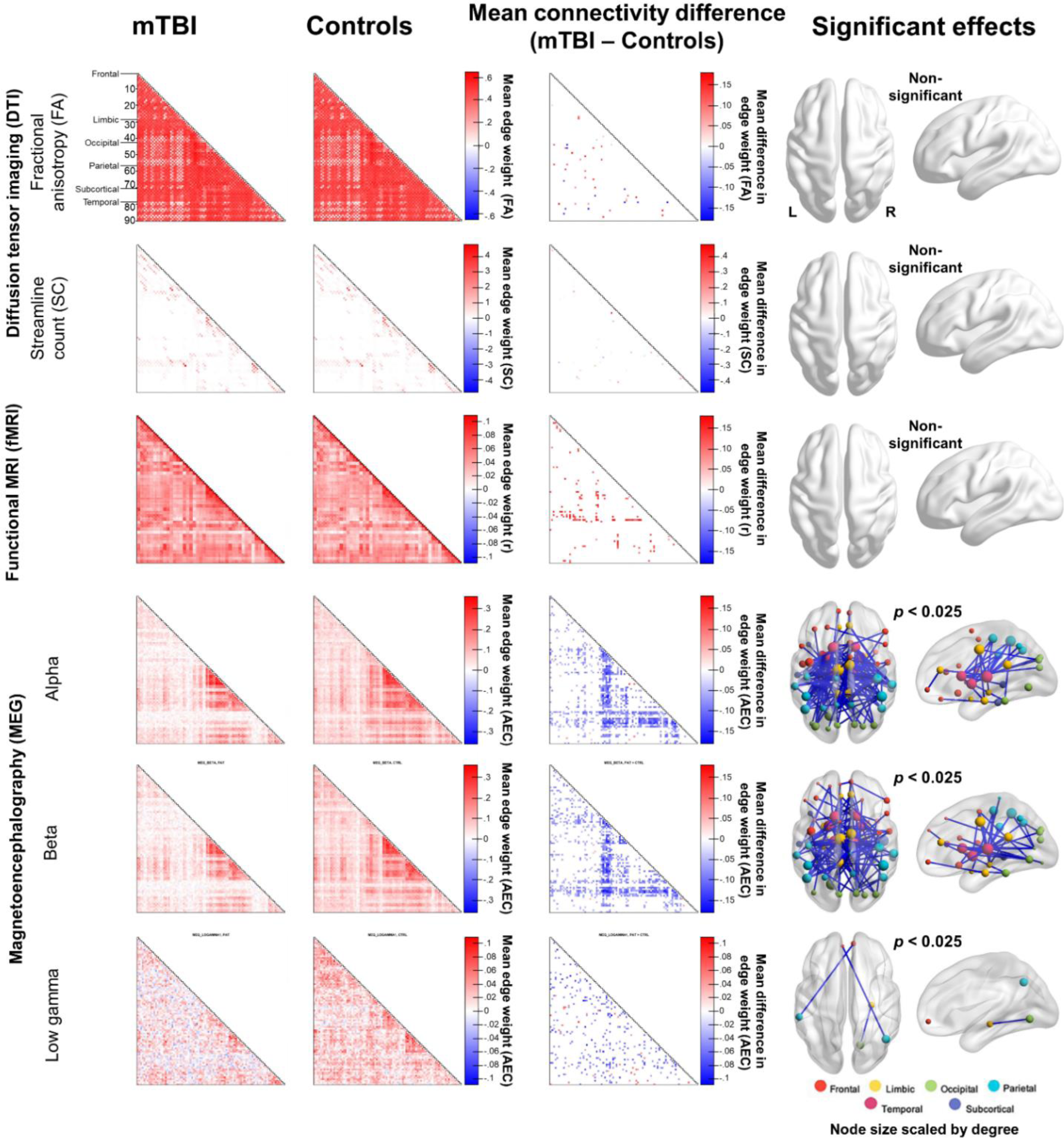
Reduced MEG functional coupling in mTBI. Reduced AEC was found in mTBI compared to healthy controls in the alpha, beta, and low gamma 1 frequency ranges (*p_corr_* < 0.025). No significant between-group effects were seen for DTI SC or FA and fMRI modalities following FDR-correction for multiple comparisons.

### Machine learning feature selection and classification analysis with single modal datasets identifying best classification performances in MEG alpha, beta, and high gamma band connectomes

The ML FS and classification modelling results contributed to the associated multimodal analysis. **Table 1** includes the AUC values from both “CV only” and “CV and holdout” processes, where the two DTI measures showed differences in ML classification modelling. Comparing the mean CV AUC value and the training data AUC value from the “CV only” analysis, the DTI FA showed signs of overfitting with the AUC on training data (0.997) substantially larger than the mean CV AUC value (0.87). The DTI SC results, however, exhibited comparable levels between the AUC on training data (0.84) and mean CV AUC (0.83).

**Table 1.** Area under the curve (AUC) values for cross validation (CV) only and CV and holdout processes.

| Dataset | CV only<br>AUC (CV<br>test data,<br>mean±SD) | CV only<br>AUC<br>(training<br>data) | CV only<br>permutation<br>test p value | CV only PLS-DA<br>permutation<br>test p value<br>(mTBI) | CV and<br>holdout AUC<br>(CV test data,<br>mean±SD) | CV and<br>holdout<br>AUC<br>(training<br>data) | CV and<br>holdout<br>AUC<br>(holdout<br>test data) |
| --- | --- | --- | --- | --- | --- | --- | --- |
| DTI FA | 0.87±0.2 | 0.997 | 0.01 | 0.001 | 0.9±0.18 | 0.993 | 0.78 |
| DTI SC | 0.83±0.25 | 0.84 | 0.01 | 0.001 | 0.92±0.14 | 0.91 | 0.78 |
| fMRI | 0.9±0.15 | 0.96 | 0.02 | 0.001 | 0.93±0.17 | 0.95 | 0.88 |
| MEG delta | 1±0 | 1 | 0.01 | 0.001 | 0.96±0.09 | 0.996 | 0.67 |
| MEG theta | 0.99±0.04 | 1 | 0.01 | 0.001 | 0.98±0.5 | 1 | 0.67 |
| MEG alpha | 0.93±0.17 | 0.98 | 0.01 | 0.001 | 0.93±0.17 | 0.998 | 0.89 |
| MEG beta | 0.98±0.08 | 0.99 | 0.01 | 0.004 | 0.94±0.1 | 0.99 | 0.89 |
| MEG low<br>gamma1 | 1±0 | 1 | 0.01 | 0.001 | 0.98±.21 | 1 | 0.67 |
| MEG low<br>gamma2 | 1±0 | 1 | 0.01 | 0.001 | 0.98±0.08 | 1 | 0.78 |
| MEG high<br>gamma | 0.98±0.05 | 0.99 | 0.01 | 0.001 | 0.98±0.08 | 0.986 | 0.89 |

For the “CV and holdout” mode results, both the DTI FA and SC datasets resulted in an AUC of 0.78 from the holdout test data. In terms of fMRI functional connectivity data, the CV models resulted in a mean AUC of 0.93, which was comparable to the AUC derived from the training data (0.95) using the final model. When classifying the holdout test data, the final fMRI SVM model exhibited an AUC value of 0.89, suggesting good performance in a close to “real world” setting. For data fusion analysis, DTI FA and SC and fMRI functional connectivity data were used.

For MEG, with comparable AUC values between the mean CV and training data performances, all seven frequencies showed minimal levels of overfitting from the “CV only” results. The performance on the holdout test data for alpha, beta and gamma was an AUC value of 0.89, suggesting good classification performance. Based on these results, the alpha, beta, and high gamma bands were used for the following data fusion analysis. **Figure 3** includes receiver operating characteristic (ROC) curves for DTI (FA, SC), fMRI, and MEG (alpha, beta, high gamma) classification performances from the “CV only” mode analysis. Group-wise heatmaps and dendrograms for each dataset are presented in **Figure S2**.

**Figure 3.**
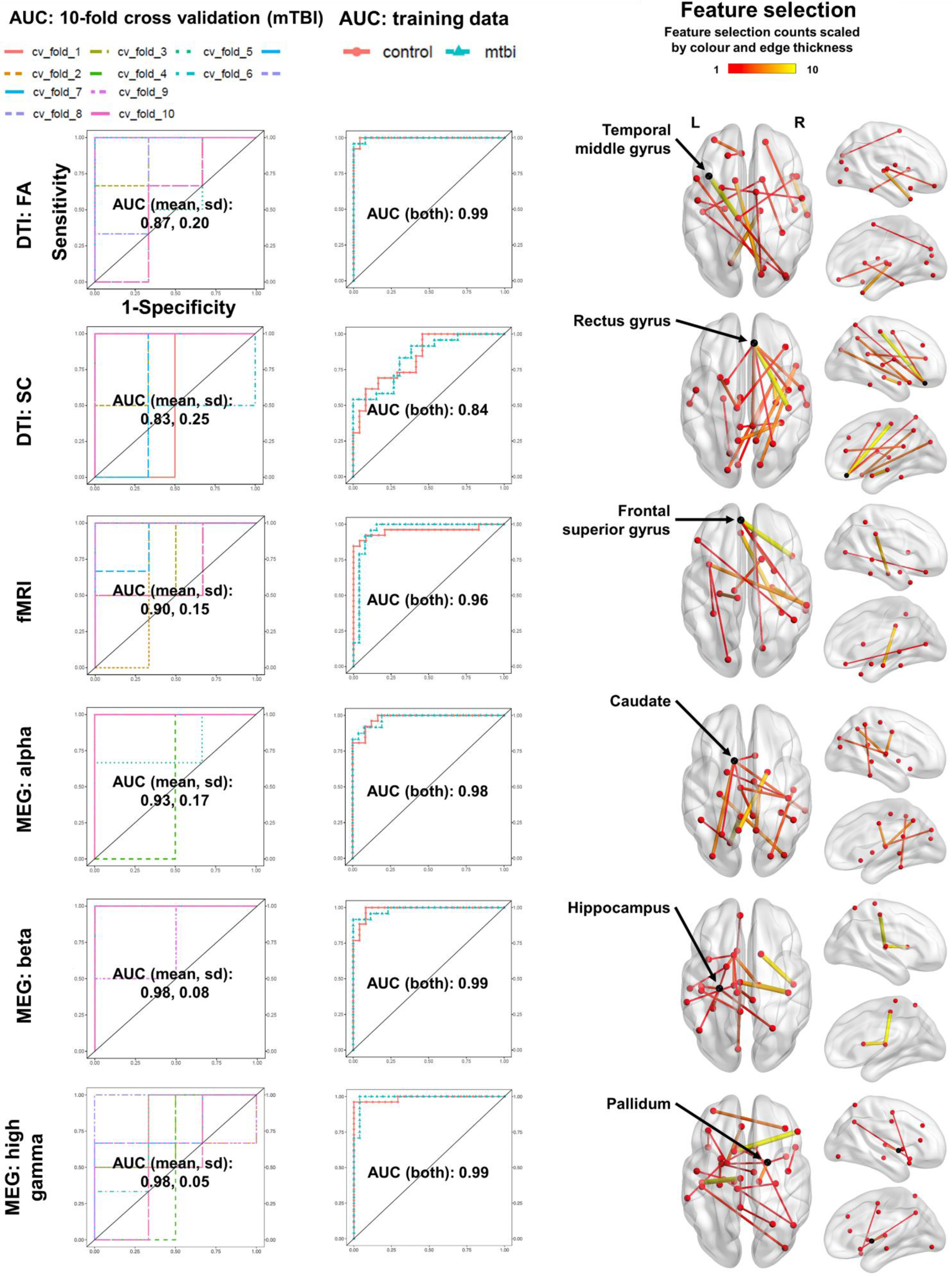
Fractional anisotropy and neurophysiological coupling provide the greatest relative discriminative accuracy for mTBI. DTI FA and MEG beta and alpha functional connectivity provide the greatest comparative accuracy for classifying mTBI from controls.

SVM and PLS-DA permutation tests were used to evaluate the features selected for DTI FA and SC, fMRI, and the seven MEG frequencies. Permutation p-values were significant (*p*<0.05) for all tests across SVM and PLS-DA algorithms (**Table 1**). The PCA score plots reveal the capacity of selected features in separating the two subject groups in an unsupervised manner (**Figure S2**). Specifically, in the subset with only the selected features, all six datasets showed a trend of group separation across the first three PCs, albeit imperfectly. These results confirmed that our ML feature selection process identified the most important features representing control and mTBI groups for each dataset. The selected features are visually represented as brain plots in **Figure 3**.

### Multimodal analysis with subject similarity fusion showing perfect group separation

To identify a “seed” dataset for fusion analysis, the single modal KNN similarity matrix was subject to hierarchical clustering analysis (heatmaps with dendrograms in **Figure 4**). Generally, the single modal hierarchical clustering on the KNN did not completely separate the mTBI subjects from the control group. Comparatively, MEG showed the best separation, with beta AEC performing best, then fMRI, then DTI. Given this finding, MEG beta was used as the “seed” dataset for the fusion analysis, where other datasets were added, to identify the best data/modality combination(s) for patient separation. Six datasets were used across the three data modalities.

**Figure 4.**
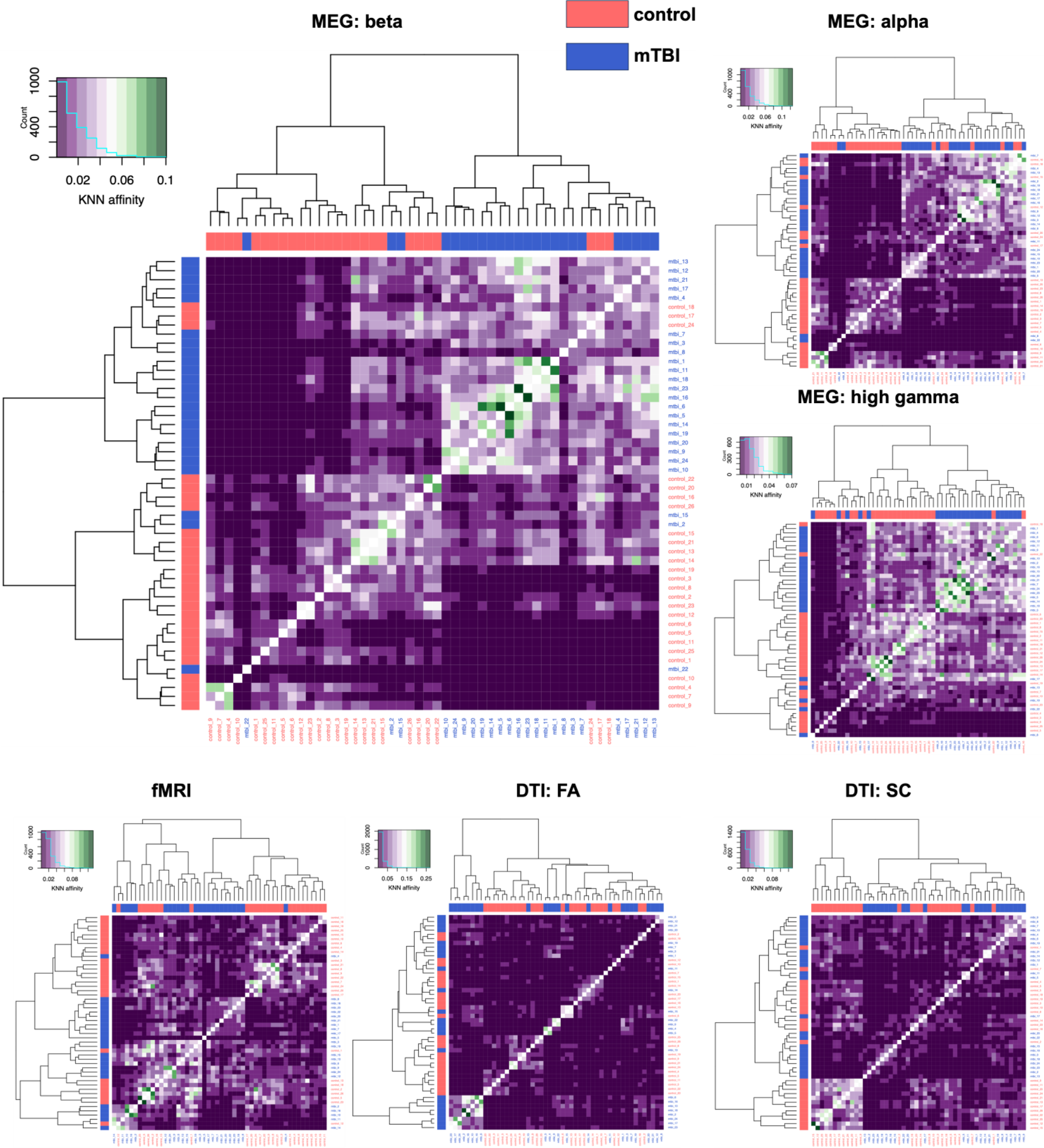
MEG beta band connectivity exhibits the best group clustering performance. Single data modality hierarchical clustering heatmaps and dendrograms show imperfect group separation.

Starting with the MEG beta KNN similarity matrix, we constructed 11 fusion networks, all featuring MEG beta matrices. We identified four fusion networks that completely stratified the groups: DTI SC+fMRI+MEG (beta), DTI SC+fMRI+MEG (alpha, beta), DTI SC+fMRI+MEG (beta, high gamma), and DTI SC+fMRI+MEG (alpha, beta, high gamma). Hierarchical heatmaps with dendrograms revealed clear two major clusters for the control and mTBI groups (**Figure 5**). The circular figures not only showed the two subject groups, but also a degree of within-group intersubject clustering. Interestingly, the MEG-only subject fusion map failed to completely separate subject groups in an unsupervised fashion.

**Figure 5.**
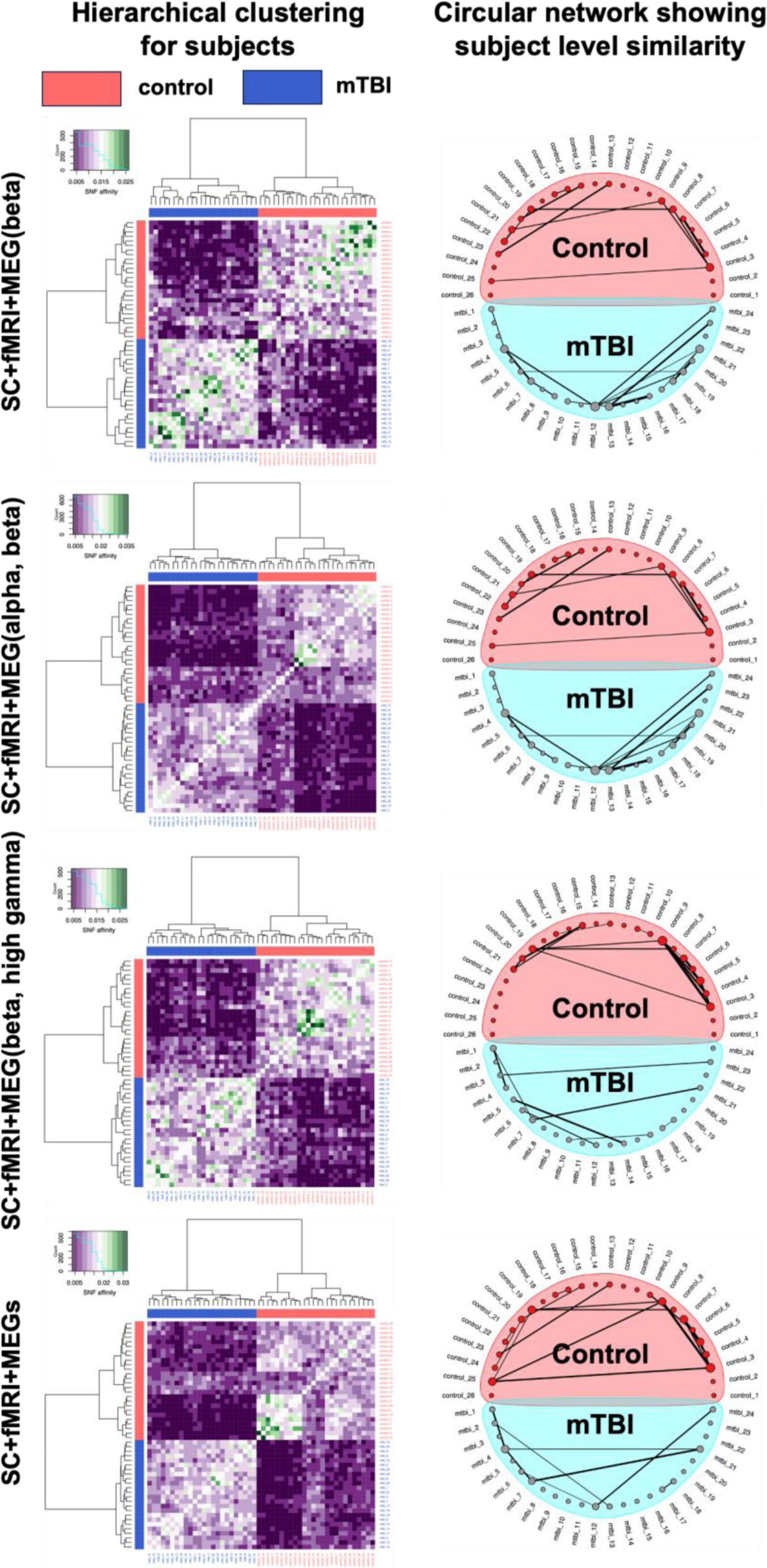
Multi data modality hierarchical clustering shows perfect group stratification. (left column) Heatmap shows various neuroimaging modality fusion combinations showing that MEG beta data led to perfect group separation, and (right column) shows subject similarity circular network plots, whose results may inform potential mTBI subtyping.

## Discussion

In the current study, we used DTI, fMRI, and MEG in individuals with mTBI to explore connectivity features that provide the best discriminative accuracy. The key findings are: 1) MEG revealed the only significant between-group effects with univariate statistics, while no group differences were found with fMRI or DTI; 2) functional connectivity (fMRI and MEG) provided the best supervised ML classification accuracy for mTBI discrimination, followed by structural connectivity (DTI), where FA outperformed streamline count; 3) supervised ML steps selected features with more connected and localized subnetworks for the DTI streamline count and MEG, than the DTI FA and fMRI connectomes, when classifying mTBI; 4) participant fusion maps featuring all three data modalities provided perfect, unsupervised separation of participants into those with and without mTBI; 5) the MEG-only full feature fusion map showed a univariate group difference for mTBI compared to non-mTBI, and this effect was eliminated upon integrating DTI and fMRI datasets.

### Neurophysiological decoupling in mTBI

Individuals with mTBI showed reduced MEG functional connectivity in the alpha, beta, and low gamma frequency ranges, with no observed effects in either DTI or fMRI, suggesting neurophysiological imaging is a sensitive approach to understand network abnormalities in mTBI. Periodic fluctuations of temporally synchronous neural oscillatory activity mediate windows for information processing and long-distance communication among brain regions, which is critical for perception, cognition, and behaviour [49], [50]. Thus, the present findings suggest disrupted long-range communication and integration of information across the brain after mTBI.

Micro-alterations in white matter tracts, namely diffuse axonal injury, are known to result from the biomechanical forces of mTBI (i.e., acceleration and deceleration; [51], [52], [53]). Diffuse axonal injury is characterized by axonal inflammation, stretching, shearing, and deafferentation, causing diffuse neurophysiological disruption to brain networks, and it is associated with chronic perceptual and cognitive sequelae [53]. Therefore, alterations to white matter tracts due to diffuse axonal injury may be a key underlying mechanism of pathophysiological coupling in mTBI.

Several studies have shown that MEG is highly sensitive to neurophysiological alterations due to brain injury in adults and youth [30], [54], [55], [56], [57]. Moreover, altered neural oscillations have been shown in those with mTBI, even in the absence of microstructural (DTI) abnormalities [55]; which suggests that MEG may be more optimal than structural imaging methods for detecting subtle effects of mTBI. The current findings provide further evidence of the value of MEG for understanding brain dysfunction in mTBI that is invisible on anatomical imaging.

This study found diminished functional coupling in the alpha, beta, and low gamma frequency bands. Reduced alpha functional connectivity has been observed in severe and mild traumatic brain injury [57], [58], [59], and our group has previously reported beta band dysconnectivity as a reliable marker of mTBI in this cohort [36]. Alpha and beta oscillations are thought to be generated in the thalamus [60], [61] and mediated by cortico-thalamic circuits [62], [63], [64] so these results suggest aberrant thalamocortical circuits in mTBI. Abnormalities in the gamma range have also been reported in adults with mTBI [56], [65], [66], [67], although there has been little research on alterations to gamma-mediated connectivity in mTBI. It is possible that decreased gamma coupling may reflect an excitatory-inhibitory (E/I) imbalance due to injury [68].

### Supervised ML analysis identifies the best performing modalities for mTBI

Using both “CV only” and “CV and holdout test” classification modelling, the supervised ML component of the multimodal integration provided performance assessment for each connectome from the three neuroimaging modalities. For the two DTI connectomes, both FA and streamline count were accurate from the “CV only” models (AUC: mean ∼0.9±SD). The “CV and holdout test” models showed a slight decrease in classification performance (AUC: ∼0.8). The discrepancy was expected as the “CV and hold out” models were trained with a portion of the data (i.e., 85% of the total subjects). These results are consistent with the previous DTI mTBI studies featuring supervised ML modelling, where proper feature selection was able to classify mTBI with promising performance [69]. Additionally, the DTI SC model was less overfit than the FA model, a finding which may be attributed to the fact that FA is sensitive to microstructural changes in brain tissue and provides more detailed information about structural connectivity than SC, making it easier for ML modelling to pick up noise or small variations in the data. Meanwhile, SC is a simpler metric representing the number of reconstructed fiber tracts, which may be less prone to overfitting as it is less specific than FA [70], [71]. For the fMRI data, both the “CV only” and “CV and holdout test” modes exhibited consistent high performance (AUC: ∼0.9) which adds to the growing evidence of effective ML based classification methods using fMRI functional connectivity [72], [73], [74]. Notably, these three aforementioned studies either required information from additional data modalities or *a priori* knowledge based feature engineering, suggesting fMRI data alone may be insufficient to effectively represent mTBI phenotypes.

A comparison of DTI and fMRI in the current study revealed that the fMRI appears to be more suitable for classification modelling with smaller sample sizes, as models trained with partial data (i.e., the “CV and holdout test” models) exhibited consistent performance with “CV only” models, which were trained with the complete data. Furthermore, when comparing classification performance between training and holdout test sets, the fMRI model exhibited a smaller drop in AUC value between the two data sets relative to the DTI models, suggesting that the fMRI model had less overfitting than the DTI models. Moreover, this finding confirms that mTBI-related functional measures may carry more sensitive signatures with greater potential for diagnostic or prognostic value relative to the subtle structural changes [75]. However, it is important to note that when resting state fMRI and DTI FA connectomes are combined we were able to classify individuals with greater performance than fMRI only [76].

In MEG, all the “CV only” models for every frequency showed accurate classification performance (AUC: mean ∼0.98±SD) with the differences among the frequency bands residing in the classification performances and level of overfitting for the “CV and holdout” models. Specifically, the alpha, beta, and high gamma bands exhibited higher performance for the “CV and hold out” models (AUC ∼0.9) than the delta, theta, and two low gamma frequency bands (AUC ∼0.7), and with lower levels of overfitting. As such, we consider alpha, beta, and high gamma bands to be the best overall performing MEG functional connectivity measures in supervised ML classification modelling, consistent with our previously reported effectiveness of beta band MEG functional connectivity in mTBI classification with the same subjects [37]. The current results further demonstrate promising performance for alpha and gamma activity, which is consistent with prior reports of functional pathophysiology in mTBI [30], [37], [57], [77], [78], [79], [80]. Similar to the fMRI-DTI comparison, MEG datasets outperformed the DTI datasets in terms of classification modelling to discriminate mTBI.

The supervised ML modelling demonstrated that functional connectivity profiles from fMRI and MEG are better than DTI structural connectivity at classifying mTBI [75]. However, prior studies have suggested that structural information may be combined with the functional imaging data to optimize ML classification performance [65], [72], [73], [74], [76]. In line with the present findings, unsupervised data integration would benefit from both structural and functional connectome data, specifically, DTI (FA or SC), fMRI, and MEG (alpha, beta, and high gamma band).

### Machine learning feature selection explores the key features driving mTBI classification

Using the connectivity features selected by the supervised ML process, we performed per participant data fusion using the six datasets across the three neuroimaging modalities identified above. To test the capability of unsupervised subject group separation with the fusion datasets, hierarchical clustering was applied to the SNF similarity scores. The MEG beta band data exhibited accurate performance when used in supervised ML based mTBI classification, consistent with our previous study [37]. Although not perfect, unsupervised hierarchical clustering also revealed that the MEG beta band provided optimal unsupervised group separation performance in discriminating individuals with mTBI from those without (**Figure 5**). Therefore, we used the MEG beta band dataset as the starting point for per participant fusion. With the MEG beta dataset as the “seed dataset,” a total of 11 dataset/modality combinations were examined, including both MEG only and DTI+fMRI+MEG participant fusion maps.

First, even with substantial group difference shown in the *full feature* fusion map (i.e., 90 x 90 maps) univariate analysis, the MEG only *subject* fusion maps (i.e., 50 x 50 maps) failed to generate complete unsupervised group separation. Indeed, only the subject fusion maps featuring all three data modalities led to perfect unsupervised subject group separation, namely DTI SC+fMRI+MEG (beta), DTI SC+fMRI+MEG (alpha, beta), DTI SC+fMRI+MEG (beta, high gamma), and DTI SC+fMRI+MEG (alpha, beta, high gamma) (**Figure 5**). Consistent with previous studies [72], [73], [74], [76], [81], [82], these results suggest that integrating both structural and functional connectivity from additional modalities (DTI and fMRI) to the seed MEG dataset substantially improves unsupervised group separation, thereby demonstrating the value of a multimodal approach to understanding injury.

Second, integrating DTI SC, fMRI, and MEG beta was sufficient to achieve complete unsupervised group separation, (i.e., without integrating additional frequency bands), which attests to the effectiveness of beta band functional connectivity stratifying mTBI patients from control subjects [36]. For DTI, only SC led to perfect subject group separation when integrated with other datasets, suggesting SC outperforms FA when separating mTBI from controls, showing that multimodal neuroimaging can inform personalized care. Indeed, multimodal neuroimaging has been explored to improve neuropsychiatric clinical practice [83].

### Limitations and Future Directions

The limitations of this study include the use of an all male cohort, and of course, given the known sex differences in mTBI for which females tend to report more mTBIs and more severe symptoms than males [84], it is necessary to include females in future work on mTBI classification using ML with data fusion, as the brain features identified for males may not be optimal for separating females with and without mTBI. Second, the age range in this study represents a young cohort (without mTBI: 20-39 years; with mTBI: 20-44 years) in which white matter tracts with fronto-temporal connections are still maturing [85]; thus, the brain features identified for separating individuals with and without mTBI in this study may not work in an older cohort with matured/aging white matter connections. Third, classification of individuals in the acute-subacute stage of mTBI was accomplished here in a cross-sectional sample. However, given that a significant minority of individuals report symptoms months after their injury [86], it would be valuable to conduct longitudinal studies to enable ML classification of those who experience persistent symptoms from those who recover. Fourth, in relation to the third limitation, the current mTBI group included both symptomatic and asymptomatic/recovered individuals and did not distinguish the former from the latter. Future work should include a larger mTBI cohort to allow for the additional classification between asymptomatic/recovered and symptomatic individuals with mTBI, both at the acute-subacute stages and in the long-term. Lastly, a relatively small sample size could be expanded the in future to increase ML model generalizability.

## Conclusions

We successfully applied ML with data fusion to determine optimal imaging modalities and brain features for separating individuals with mTBI from those without. Notably, univariate group differences were only found for functional connectivity, as quantified by MEG, with no mTBI-related distinctions identified by either fMRI or DTI datasets, lending further evidence to the advantageous sensitivity of MEG to brain dysfunction in mTBI that is otherwise invisible to fMRI and structural MRI. The combined assessment of functional connectivity by MEG and fMRI exhibited robust unimodal classification accuracy for mTBI, followed by structural connectivity (DTI), for which FA provided sightly superior classification performance relative to SC; however, SC outperformed FA for data interpretation and fusion. Ultimately, integration of all three data modalities allowed for perfect, unsupervised separation of participants with and without mTBI, and although the MEG-only full feature fusion map showed group differences, addition of the DTI and fMRI datasets removed this effect. The brain markers identified by ML with data fusion in this study corroborate previous multimodal investigations in mTBI and highlight the importance of considering modality-specific factors in understanding network abnormalities associated with mTBI.

## Data Availability

All data produced in the present study were funded through defence contracts, and given the nature of the work, are not publicly available.

