## Supplemental Materials for "The structural, functional, and neurophysiological connectome of mild traumatic brain injury: a DTI, fMRI and MEG multimodal clustering and data fusion study"

### Supplementary Methods

*MEG methods*

Artefacts related to cardiac activity, eye blinks, and eye movements were removed using the independent component analysis (ICA) function ('fastica') in Fieldtrip. Components were visually inspected and rejected by an experienced MEG analyst. As many epochs of 10s as possible were selected from the 5 min recording such that: (a) the head position during each epoch deviated less than 8 mm from the recording median, and (b) excluded portions of data that contained SQUID resets or exceeding a threshold of ±2 pT after ICA component rejection. A maximum of 24 artefact-free trials were selected, with the mTBI group having a mean of 21.8 trials (SD +/-6.1), and the control group a mean of 22.8 trials (SD +/- 3.5), with no significant difference 210 between the groups in remaining number of trials ($t(48)=-0.7$, $p=0.49$).

For MEG data co-registration, a single-shell head model for each participant was generated based on a their anatomical T1-weighted MRI image. A beamformer was used to recover time series from 90 regions of the Automated Anatomical Labelling (AAL) atlas [1]. The centroid of each AAL parcel was used to define the node location for the beamformer reconstruction. The Fieldtrip implementation of the linearly constrained minimum variance (LCMV) vector beamformer [2] was used to reconstruct the neural time series at each location defined by the AAL centroid, with 5% Tikhonov regularization. Lead fields were computed from the template single shell head model for a unit current dipole in 3 dimensions at each node. The beamformer weights for the activity at each node were computed by projecting the sensor weights along the axis with the highest singular value decomposition (SVD) variance resulting in a one-dimensional activity time series for each node.

The time series data retained from the virtual electrodes were z-scored (i.e., mean centred, variance normalised). The broadband regional time course for each node location was filtered into delta (1-3 Hz), theta (3-7 Hz), alpha (8-14 Hz), beta (15-25 Hz), low gamma 1 (30-55 Hz), low gamma 2 (65-80 Hz), and high gamma (80-150 Hz) ranges. A symmetric orthogonalization leakage correction procedure was applied to the filtered regional time series to attenuate artificially inflated coupling that might be the result of beamformer leakage. The Hilbert transform was applied to the resultant time course to derive instantaneous estimates of the amplitude envelope, which was then down-sampled to 1 Hz by averaging the amplitude over 1s intervals, similar to other studies of MEG functional coupling [3]. Pearson correlations between all node pairs were calculated to index functional coupling. AEC was chosen over other measures of neural communication (for example, in contrast to the phase lag index or phase locking value) as it has been shown to be the most reliable measure of connectivity across sessions and over individuals, pointing to the greatest replicability as well as the lowest susceptibility to co-registration related errors [4].

*fMRI methods*

Before preprocessing, the first 5 volumes of the functional images for each subject were removed to allow for optimal steady-state magnetization to be achieved as well as accounting for participant adaptation to scan environment. Functional image preprocessing was carried out using the Data Processing Assistant for Resting-State fMRI (DPARSF) toolbox [5] and the SPM8 package (SPM, http://www.fil.ion.ucl.ac.uk/spm), employing standard preprocessing steps. Slice time correction was performed for the interleaved sequence acquisition, and realignment of the fMRI BOLD data was done by calculating a six-parameter rigid body spatial transformation. [6] Since none of the participants exhibited more than 3 degrees rotation and 3 mm displacement in any direction for the duration of the scans, no volumes were discarded. Normalization was performed using unified T1 segmentation onto the Montreal Neurological Institute (MNI) space template in SPM8. Smoothing was performed using a Gaussian kernel at FWHM = 7 mm after all images were resampled to 3.5 mm isotropic voxels. The global signal was removed along with nuisance covariance regressors such as head motion parameters, the white matter signal (WM), and the cerebrospinal fluid (CSF) in order to minimize effects of non-neuronal oscillations and motion [7], [8], [9], [10]. Images were band-pass filtered between 0.01- 0.1 Hz to retain low frequency oscillations in the resting state fMRI which are believed to reflect neuronal activity [11] while suppressing low frequency drifts and high frequency noise resulting from other psychological activities [12], [13].

Brain network average time domain signals were extracted from 90 cerebral regions, again as defined by the automated anatomical labeling (AAL) atlas provided by the Montreal Neurological Institute. [1] Mean time series signals were calculated by averaging the voxel time courses within each of the 90 regions which served as a node in the network. Regional correlation matrices were calculated for each subject (for each node pair) based on the Pearson's correlation coefficient between the time-courses, creating the edge weight for the network.

*Diffusion Tensor Imaging*

DTI Images were preprocessed using the fMRI Software Library (FSL, v. 6.0.1) and Mrtrix (v. 3.0) [14]. Gibbs Ringing Removal [15] followed by PCA denoising [16] was performed first in MRtrix3 [14].

EDDY [17] with outlier detection [18] was used to correct for eddy current induced distortions as well as susceptibility-induced distortions. Data were brain extracted using BET [19]. For each subject, the first volume in each diffusion data set (b = 0) was aligned with its corresponding T1 anatomical image using a linear registration in FSL (FLIRT). [20] The anatomical image for each subject was then aligned with MNI space using a linear transformation (FLIRT) followed by a non-linear transformation (FNIRT) [21] in FSL. The AAL-90 atlas parcellations [1] were then transformed into the anatomical space of each individual subject by inverting the preceding non-linear transformations. Finally, the parcellations were brought into diffusion space using the inverse transformation matrix from the b0 image to anatomical image registration. These parcellations were later utilized to inform connectome construction.

Response functions for single-fibre WM as well as GM and CSF were estimated from the data themselves using an unsupervised method [22]. Single-Shell 3-Tissue Constrained Spherical Deconvolution (SS3T-CSD) [23] was performed to obtain WM-like Fiber Orientation Distributions (FODs) as well as GM-like and CSF-like compartments in all voxels using MRtrix3Tissue (https://3tissue.github.io/). Following SS3T-CSD, 3-tissue bias field and intensity normalisation was performed [24] followed by filtering of tractograms [25]. Anatomically Constrained Tractography (ACT) [26] using a five-tissue-type (5TT) segmented image was utilized along with the previously registered AAL-90 parcellations to generate single subject connectomes quantified by Streamline Count (SC) and Fractional Anisotropy (FA).

*Supervised machine learning based feature selection*

For feature selection, a recursive random forest feature selection and SVM modelling assessment were featured during the CV iterations (CV-rRF-FS-SVM). For each feature selection iteration, a list of features was selected by rRF-FS and evaluated by SVM. With the 10 feature lists from the 10-fold CV-rRF-FS-SVM selection, a voting process was used to select top count features. Specifically, features selected at least two times were retained for the final consensus list of features.

The final consensus features were then used to build a final SVM model through another 10-fold CV process. We used the “linear” kernel for SVM modelling steps, accordingly to our preliminary tests (data not shown). First, the final SVM model was used to determine the effectiveness of the selected features in classifying the two subject groups through a permutation test (99 iterations). For the permutation test, the sample labels were randomly shuffled to build permutation CV models through the same 10-fold CV process (used to build the final CV model). The mean CV AUC values were then compared between the final and permutation SVM models. A permutation p-value was calculated for the permutation test to assess if the final SVM model significantly outperformed the permutation SVM models in classification AUC. Both the permutation test and p-value calculation followed Ojala and Garriga [27]. Second, the performance assessment was conducted for the final SVM model. Specifically, the mean CV AUC (±SD) was reported as the final performance for the "CV only" mode process. For the "CV and holdout" mode, the final performance was defined by the classification performance (AUC) of the holdout test data when applied with the final SVM model.

Additionally, partial least squares-discrimination analysis (PLS-DA) was used as a separate classifier to test the classification performance of the selected features beyond SVM. This step was used to assess the classification versatility of these features, and in turn, to further confirm the effectiveness of our ML feature selection method. To determine performance, the same permutation test and p-values were used on the PLS-DA models (999 iterations).

*Supervised machine learning implementation*

The supervised ML component was used to build classification models, which were subsequently used to determine datasets with the best classification performance for integration steps. Further, the supervised ML workflow included a feature selection functionality that was used to extract most relevant features in subject group differentiation from each data modality. The selected features were used for subject fusion and modality correlation analyses.

The supervised ML analysis was conducted in two modes: "cross validation only (or CV only)" and "cross validation with holdout test (or CV and holdout)". The major difference between the two modes resides in the data resampling process. For the "CV only" mode, the entire subject list was used for the 10-fold random data split for both feature selection and classification modelling steps. The "CV and holdout" mode, however, randomly drew 15% of the subjects as a holdout test set, with the 85% of the data going through the same feature selection and classification process as the "CV only" mode. All random data resampling steps used a stratified approach, ensuring all data folds or partitions featured the same subject group ratio.

To ensures all available data were used for feature selection, the "CV only" mode was used for feature selection. As such, the subsequent multimodal analysis used the features selected from this mode. The "CV and holdout" mode, meanwhile, tests the close to "real world performance" in mTBI classification with single modality data. Therefore, results from both the "CV only" and "CV and holdout" modes were used to determine classification modelling performance for the datasets/modalities. Specifically, "CV only" mode results were used to identify the least overfitted frequency bands, while the "CV and holdout" mode was used to identify the frequency band leading to the best “real world” classification performance. Ultimately, the frequency bands exhibiting the least overfitting and the best “real world” classification performance were retained for the multimodal integration analysis.

*Multimodal fusion analysis*

For all the datasets, SNF-based data fusion was carried out using k-nearest neighbour (KNN) affinity matrices [28]. For subject fusion, a KNN similarity matrix was built for each dataset as a normalization step, using the respective initial connectivity matrices, and SNF fusion was conducted using the KNN similarity matrices. Regarding the subject similarity fusion, the KNN similarities were derived from the Euclidean distance matrices (the SNF default input) of only the ML selected features. All data were centered and normalized prior to KNN and SNF processes.

The KNN similarity matrix computation was the process of normalizing and transforming the input data matrices according to the neighbour number threshold. The process resulted in two matrices for each data type: a “P” matrix containing full local and remote similarity information, and an “S” matrix that only included local similarity. The local similarity was determined by the mean local connection for each element of the matrices up to the neighbour number threshold. For the data fusion, briefly, the SNF step followed the message-passing theory [29] where each network was updated iteratively with the information from each other until they converge into a single network. The data fusion used the “P” matrix as the initial state and the “S” matrix as the kernel for each iterative update between data modalities during fusion. The “P” matrices were calculated using with the “affinityMatrix” function from the SNFtool R package [28]. The default settings were used, with K=20 (number of neighbours) and sigma=0.5, whereas the “S” matrices were computed during the data fusion. The SNF process was carried out using the “SNF” function from the SNFtool package with default settings, t=10 and K=20. The detailed mathematical representation of the entire process can be viewed in the original publication [28].

For subject fusion analysis, dendrogram associated heatmaps and circular network views were used as visual representations of the results. For heatmaps, the dendrograms showed the hierarchical clustering results, while the colour-coded heatmap represented either KNN or SNF similarity depending on the nature of the network (i.e., single or multi-modality). Similarly, in addition to clustering pattern (node colour) and similarity (edge thickness), the circular networks depicted additional information, including inner and cross cluster connectivity (edge colour) and most connected (highest degree) nodes (node size). For the subject similarity networks, we identified and displayed the nodes (per major cluster) passing the top 5 percentile of all node degrees, as well as edges passing the top 2 percentile of all similarities. Regarding the number of hierarchical clusters, we manually chose to identify two clusters for the subject similarity analysis, as per two subject groups. This way, we were able to assess if the subjects were correctly categorized into their respective groups.

The same permutation based ANCOVA analysis (with age as a covariate) was conducted for the full feature fusion maps comparing between concussion and healthy control groups, featuring the dataset/modality combinations for the subject fusion maps with perfect unsupervised subject separation.

#### Supplementary Results

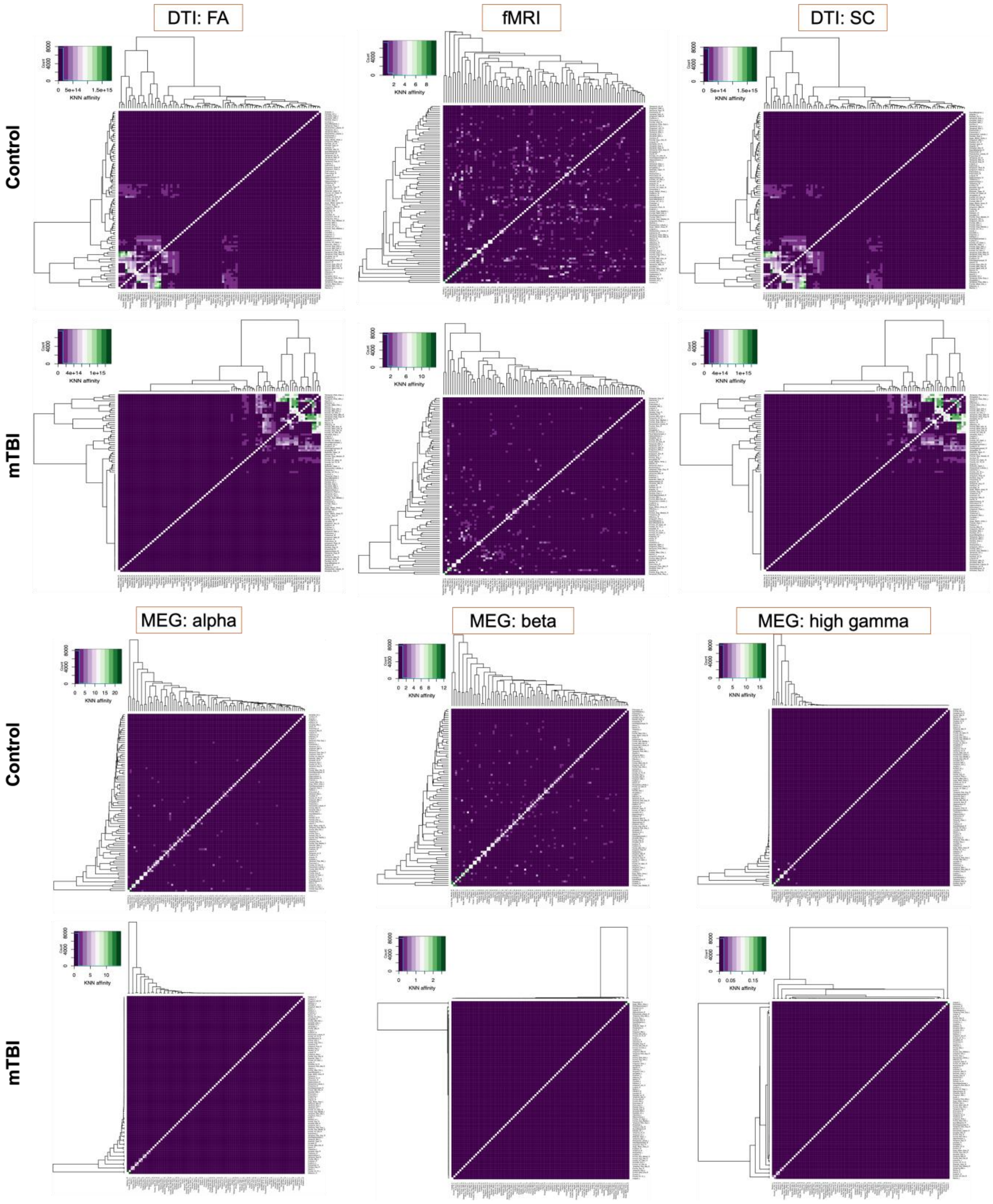


**Fig S1. Group-wise heatmaps and dendrograms for each dataset.**

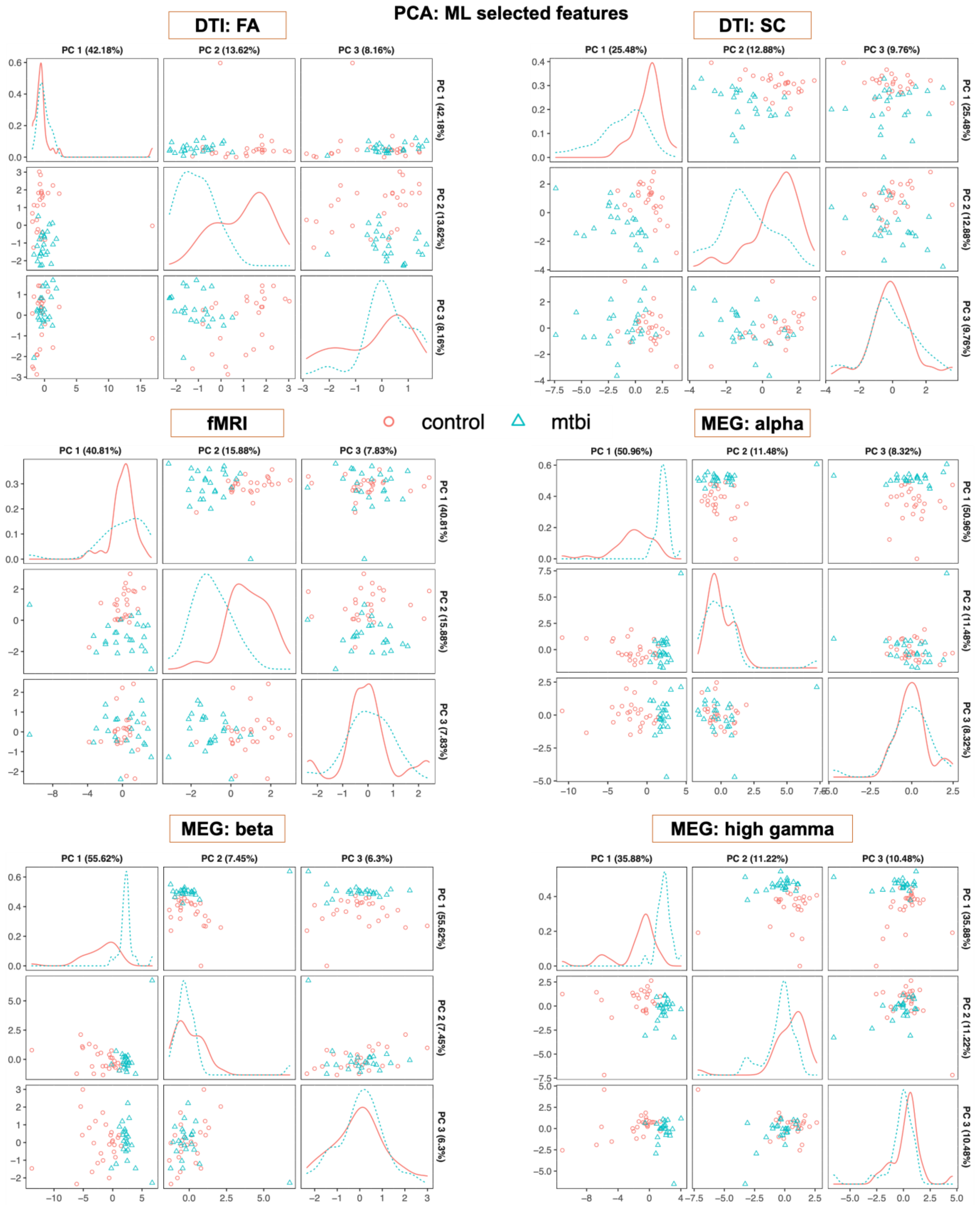


**Figure S2. Machine learning (ML) selected features.**

[22] T. Dhollander, R. Mito, D. Raffelt, and A. Connelly, "Improved white matter response function

estimation for 3-tissue constrained spherical deconvolution," *Proc. Intl. Soc. Mag. Reson. Med*, no. May 11-16, p. 555, 2019, [Online]. Available: https://www.researchgate.net/publication/331165168_Improved_white_matter_response_function_estimation_for_3-tissue_constrained_spherical_deconvolution

[29] J. Pearl, *Probabilistic Reasoning in Intelligent Systems: Networks of Plausible Inference*. San Francisco, CA, USA: Morgan Kaufmann Publishers Inc., 1988.
